# Integrating Social Determinants of Health with SOFA Scoring to Enhance Mortality Prediction in Septic Patients: A Multidimensional Prognostic Model

**DOI:** 10.1101/2024.03.13.24304233

**Authors:** Elie Sarraf, Alireza Vafaei Sadr, Vida Abedi, Anthony S Bonavia

## Abstract

**Background:** The Sequential Organ Failure Assessment (SOFA) score is an established tool for monitoring organ failure and defining sepsis. However, its predictive power for sepsis mortality may not account for the full spectrum of influential factors. Recent literature highlights the potential impact of socioeconomic and demographic factors on sepsis outcomes.

**Objective:** This study assessed the prognostic value of SOFA scores relative to demographic and social health determinants in predicting sepsis mortality, and evaluated whether a combined model enhances predictive accuracy.

**Methods:** We utilized the Medical Information Mart for Intensive Care (MIMIC)-IV database for retrospective data and the Penn State Health (PSH) cohort for prospective external validation. SOFA scores, social/demographic data, and the Charlson Comorbidity Index were used to train a Random Forest model using the MIMIC-IV dataset, and then to externally validate it using the PSH dataset.

**Findings:** Of 32,970 sepsis patients in the MIMIC-IV dataset, 6,824 (20.7%) died within 30 days. The model incorporating demographic, socioeconomic, and comorbidity data with SOFA scores showed improved predictive accuracy over SOFA parameters alone. Day 2 SOFA components were highly predictive, with additional factors like age, weight, and comorbidity enhancing prognostic precision. External validation demonstrated consistency in the model’s performance, with delta SOFA between days 1 and 3 emerging as a strong mortality predictor.

**Conclusion:** Integrating patient-specific information with clinical measures significantly enhances the predictive accuracy for sepsis mortality. Our findings suggest the need for a multidimensional prognostic framework, considering both clinical and non-clinical patient information for a more accurate sepsis outcome prediction.

## INTRODUCTION

The diagnosis of sepsis has long relied on measures of organ dysfunction such as the Sequential (or Sepsis-Associated) Organ Failure Assessment (SOFA)[1]. Established in 1996 to monitor organ failure across a spectrum of critical illnesses, a change in the SOFA score of ≥2 points, in the context of infection, currently defines sepsis [1, 2]. Despite its widespread use, the SOFA score was originally based on a general scale of illness severity and was not sepsis-specific. Other severity-of-illness scores exist, although no scoring system to date is tailored exclusively to predicting mortality in sepsis [3]. Additionally, these scores are predominantly based on laboratory data, which may not keep pace with the clinical condition of critically ill patients. Among the six SOFA domains, four hinge on laboratory results, potentially delaying the identification of critical changes in a patient’s condition.

Recent literature suggests that socioeconomic status and demographic background may play a more definitive role in the outcomes of sepsis than previously recognized [4-6]. In fact, artificial intelligence has facilitated data-driven approaches to uncovering existing healthcare disparities [7]. These social determinants of health may provide immediate, specific insight into patient risk without the need for expensive tests. Thus, our study sought to assess the predictive power of SOFA scores relative to demographic and socioeconomic factors for sepsis prognosis. We hypothesized that a combined approach using SOFA scores measured on the days following sepsis onset, together with social health determinants, could offer a more accurate prediction of mortality.

## METHODS

We accessed anonymized patient data from the Medical Information Mart for Intensive Care (MIMIC)-IV database (version 2.2, Jan 6 2023), which includes critical care data from Beth Deaconess Medical Center [8]. This resource was chosen for its comprehensive data and patient mortality information. Using existing code [9], we calculated SOFA scores for patients fitting Sepsis-3 criteria [1].

From the onset of suspected infection, we tracked the highest daily SOFA score and its change between each day of critical illness. Additionally, we computed daily changes in SOFA (delta SOFA) and cumulative daily SOFA scores (sum SOFA). This method ensured data completeness, with no need for imputation except when patients left the ICU. We also gathered demographic data and the Charlson Comorbidity Index (CCI, [10]), focusing on 30-day mortality as the primary outcome.

Using Python’s *scikit-learn* library (version 1.3.2)[11], we trained a Random Forest model, evaluating it through 5-fold cross-validation. We tested six model types, including (1) models that were trained on 3-day and 8-day retrospective data, (2) models with and without daily SOFA organ component measures, and (3) models with and without patient socioeconomic, demographic and comorbidity data. Metrics such as feature importance, PPV, NPV, sensitivity, specificity, and AUROC were assessed during each iteration.

We then externally validated our best predictive model with real-world, observational data from critically ill patients having sepsis and forming part of a prospective research cohort at Penn State Health (PSH) from Aug 2020 – Feb 2024. Given that the validation data set included SOFA scores measured every other day, the original model was re-tuned to only use odd-numbered days. We also compared the featured importance between a streamlined dataset for a Random Forest model with both the MIMIC and PSH data. Our analysis code is available on Github, and the study adhered to TRIPOD guidelines for predictive model reporting [12].

## RESULTS

Of 299712 unique patients included in the MIMIC-IV database, 32970 had sepsis. Among these, 57.8% were male and the mean age was 66.7 ± 16 years. A total of 6824 patients (20.7%) died within 30 days of the sepsis onset.

For each of the six scenarios described above, 25 different machine learning models were trained. **Figure 1A** describes five metrics pertaining to the model having the highest comparative AUROC (75%, with 95% CI 73 – 77%). This model demonstrated 77% sensitivity (95% CI 75 - 77%), 74% accuracy (95% CI 72 - 76%) and a precision of 43% (95% CI 39 – 47%). The most effective model combined demographic, socioeconomic, and comorbidity data with total SOFA score and SOFA organ component measures over an 8-day period post-sepsis. However, adding just demographic, socioeconomic, and comorbidity data to the total SOFA score (i.e., without individual organ dysfunction measures) significantly enhanced 30-day mortality prediction as compared with total SOFA score alone. This improvement was evident in analyses conducted at both 3 and 8 days after sepsis onset.

**Figure 1.**
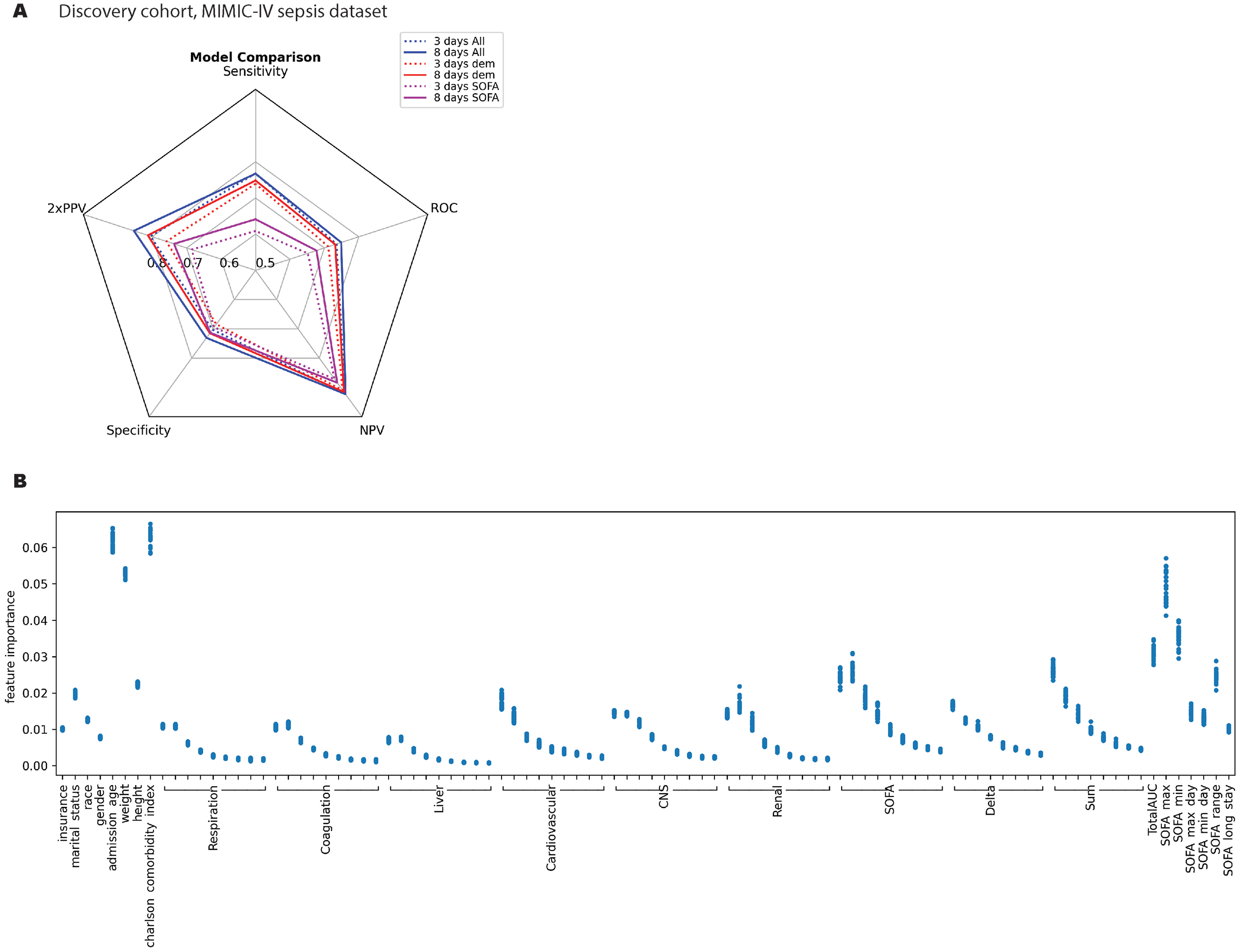
Comparative Metrics and Feature Importance for 30-Day Mortality Prediction Based on Parameters derived from Medical Information Mart for Intensive Care (MIMIC)-IV dataset. (A) This spider plot illustrates the efficacy of different predictive models over a span of 3 and 8 days following a sepsis diagnosis. ‘SOFA’ represents models utilizing exclusively SOFA-related parameters, including total SOFA score, its daily change (delta), cumulative sum, and AUROC values. ‘Dem’ extends the ‘SOFA’ model by incorporating demographic and socioeconomic factors such as insurance status, marital status, ethnicity, age, gender, body weight, height, and the Charlson comorbidity index. ‘All’ encompasses all ‘Dem’ variables plus daily-specific organ function scores (e.g., day-to-day SOFA respiratory scores). Metrics of model performance include positive predictive value (PPV), negative predictive value (NPV), and the receiver operating characteristic (ROC) curve area. (B) This figure displays the relative feature importance of various parameters in predicting 30-day mortality following sepsis. The x-axis is segmented by square brackets indicating an 8-day timeline post-sepsis onset, with day 1 at the bracket’s left end progressing to day 8 at the right. The parameters include daily SOFA component scores for respiration, coagulation, liver function, cardiovascular stability, central nervous system (CNS) activity, and renal performance. ‘Delta SOFA’ quantifies the day-over-day variation in SOFA scores, while ‘sum SOFA’ aggregates SOFA scores over two consecutive days. ‘Total AUC’ represents the cumulative SOFA score up to the current day. ‘SOFA max’ and ‘SOFA max day’ denote the peak SOFA score recorded for a patient and the specific day it was registered. Conversely, ‘SOFA min’ and ‘SOFA min day’ indicate the lowest SOFA score and its corresponding day. ‘SOFA range’ is the time span between the highest and lowest SOFA scores, providing a measure of fluctuation in organ function. ‘SOFA long stay’ tallies the total days a patient’s SOFA scores are monitored within the ICU, reflecting the duration of critical care received.

With respect to feature importance, SOFA organ component scores measured on day 2 were most predictive of 30-day mortality (**Figure 1B**). Amongst organ systems affected by sepsis, cardiovascular dysfunction, renal dysfunction, and central nervous system dysfunction were most predictive of 30-day mortality. Equally important was the relative feature importance of age, weight, height, marital status and CCI as compared with organ-specific or overall SOFA measures (**Figure 1B**).

**Figure 2A** illustrates the results of external validation using prospective data from 105 septic, critically ill patients undergoing care at PSH. In this data set, 52.4% of patients were male, with a mean age was 66.5 ± 15 years. Nineteen of these patients died within 30 days. When individual metrics were compared between discovery and validation cohorts, there was a noted increase in sensitivity and negative predictive value, and to a minor degree AUROC, at the expense of a lower specificity and positive predictive value. The negative predictive value of the model derived from MIMIC data appeared to decrease when using data derived from every other day of critical illness. Otherwise, the model’s performance remained consistent.

**Figure 2.**
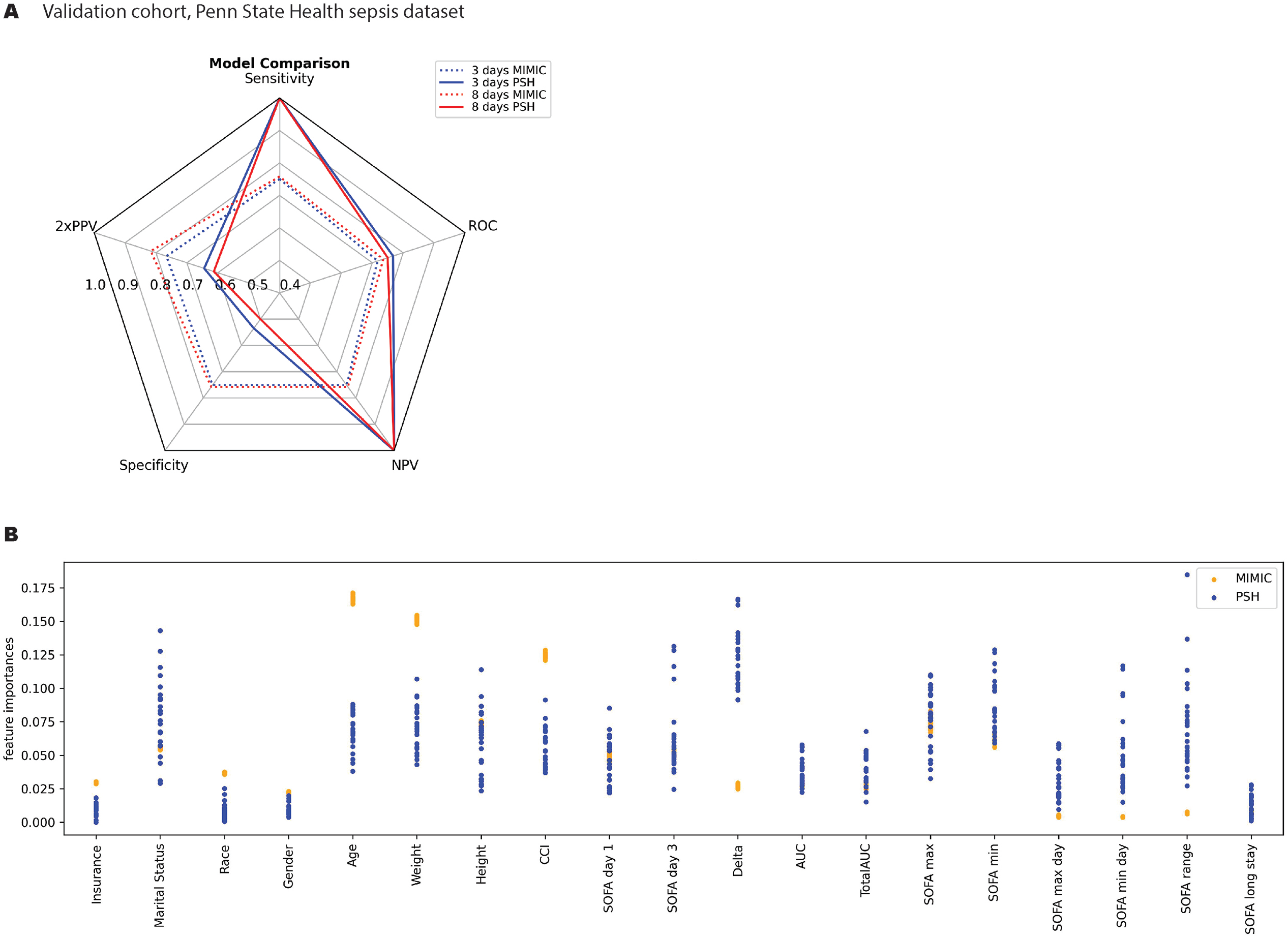
External Validation of 30-Day Mortality Prediction Model, Utilizing Data from Septic Patients Treated at Penn State Health (A) This spider plot compares the performance of the predictive model trained on the MIMIC dataset on a validation subset of MIMIC (‘MIMIC’) versus the external validation data obtained from Penn State Electronic Medical Record (‘PSH’). All abbreviations are defined in the legend for Figure 1. (B) This figure compares the relative feature importance in predicting 30-day mortality following sepsis for MIMIC (blue) and PSH (orange). We extracted patient characteristics and clinical variables from the first three days of illness onset alone. To minimize the risk of model overfitting, we streamlined the number of features following the initial discovery phase.

Interestingly, in the streamlined model, age, weight, height and CCI continued to comprise the most important features (**Figure 2B**). The importance of marital status was variable as compared with results derived from the MIMIC model, while ‘delta SOFA’ between days 1 – 3 of critical illness proved to be far better correlated with mortality in the PSH data set.

## DISCUSSION

The cornerstone of sepsis diagnosis has traditionally hinged on the monitoring of organ dysfunction via the SOFA score [1]. Historically, a higher SOFA score, signaling severe organ failure, has been closely linked with poorer clinical outcomes [13, 14]. However, our analysis proposes a paradigm shift. Our pivotal finding is the pronounced impact of patient-specific and social risk factors on 30-day mortality, overshadowing the predictive relevance of organ dysfunction severity.

Our data indicate that, by day 2 of sepsis, certain organ dysfunction measures can predict mortality with some degree of reliability. Yet, it is the integration of a patient’s age, weight, height, marital status and comorbidity profile (as encapsulated by the Charlson Comorbidity Index) that amplifies the prognostic precision significantly more than the SOFA score alone or any of its individual organ components. By Day 8, models enriched with these variables not only sustained but also enhanced the predictive accuracy of mortality, with model performance metrics surpassing those based solely on clinical measures.

The inclusion of individual organ dysfunction parameters only modestly improved the model’s performance, reinforcing the premise that, while clinical measures of organ dysfunction are not to be overlooked, they are evidently less predictive of patient outcomes compared to social determinants of health. This is a critical observation, as it underscores the limitations of current clinical-only prognostic models and highlights the potential for improved risk stratification through the incorporation of other patient-specific factors.

We externally validated the best model generated from publicly available data on a subset of patients receiving health care at our institution. We found that, while the predictive model remained stable, delta SOFA between days 1 and 3 of critical illness was a far stronger predictor of mortality in our patient population. The prognostic utility of delta SOFA has been previously reported [15], although it is not currently in widespread clinical use.

Past efforts to improve on the SOFA score have produced mixed results [16-18]. However, our research supports these endeavors, suggesting that an enhanced SOFA score incorporating social determinants of health could sharpen the predictive accuracy for mortality in sepsis patients. This augmented model may be particularly beneficial for patients who, due to cognitive impairments caused by their illness, are unable to provide a detailed medical history.

Although our study offers valuable insights, it is important to acknowledge its limitations. A significant constraint was that our validation dataset recorded SOFA scores every 48 hours instead of every 24 hours. This less frequent recording could have affected the validation’s accuracy, given that peak SOFA scores usually manifest within the first 24 hours of sepsis onset. Furthermore, our exclusive use of the Sepsis-3 diagnostic criteria might not encompass the full range of clinical presentations and outcomes. This is particularly relevant since other frameworks, like Sepsis-2 [19], are still widely used in clinical practice.

In conclusion, our investigation underscores the imperative to revisit the prognostic frameworks for sepsis. It is evident that a multidimensional approach, encompassing both clinical and non-clinical factors, is crucial for a more accurate prediction of outcomes. Our work contributes to the growing body of evidence that supports the integration of broader patient information, extending beyond the confines of physiological and laboratory measures, into prognostic models for sepsis.

## Data Availability

All data produced in the present study are available upon reasonable request to the authors

## List of abbreviations

AUROC: area under the receiver operating characteristic curve
CCI: Charlson comorbidity index
ICU: intensive care unit
MIMIC: Medical Information Mart for Intensive Care
NPV: negative predictive value
PPV: positive predictive value
PSH: Penn State Health
SOFA: sequential (or sepsis-associated) organ failure assessment

## DECLARATIONS

### Ethics approval and consent to participate

De-identified and publicly-available human data was accessed via Physionet [20] and utilized in accordance with PhysioNet Credentialed Health Data Use Agreement 1.5.0. Prospective validation data was collected in accordance with the Penn State Human Subjects Protection Office (IRB study# 15328, approved 7/27/2020).

### Consent for publication

Not applicable.

### Availability of data and materials

The datasets analyzed during the current study are available in the MIMIC-IV version 2.2 database repository, available at https://physionet.org/content/mimiciv/2.2. Validation data from PSH will be made available from the investigator on reasonable request. The code used to analyze MIMIC-IV data is publicly available and can be accessed here: https://github.com/rasman/SOFA.

### Competing interests

The authors declare that they have no competing interests.

### Funding

Funding was provided by the National Institute of General Medical Sciences R35GM150695 (ASB), endowment funds from the Department of Anesthesiology at Penn State Health (ES), and start-up funds from the Department of Public Health Sciences (VA).

### Authors’ contributions

ES contributed to data curation and preprocessing. ES and AVS were responsible for the development and implementation of the machine learning algorithms and performed the data analysis. VA provided critical feedback, and helped shape the research, analysis, and manuscript. ASB conceptualized the study, assisted in the interpretation of the computational data analysis, supervised the project, and led manuscript writing. All authors read and approved the final manuscript.

## Acknowledgements

Not applicable.

## Notes

### Competing Interest Statement

The authors have declared no competing interest.

### Funding Statement

This study was funded by the National Institute of General Medical Sciences

### Author Declarations

Ethics committee/IRB of Penn State University gave ethical approval for this work

## REFERENCES

1. Singer M, Deutschman CS, Seymour CW, Shankar-Hari M, Annane D, Bauer M, Bellomo R, Bernard GR, Chiche JD, Coopersmith CM et al: The Third International Consensus Definitions for Sepsis and Septic Shock (Sepsis-3). JAMA 2016, 315(8):801–810.

2. Vincent JL, Moreno R, Takala J, Willatts S, De Mendonca A, Bruining H, Reinhart CK, Suter PM, Thijs LG: The SOFA (Sepsis-related Organ Failure Assessment) score to describe organ dysfunction/failure. On behalf of the Working Group on Sepsis-Related Problems of the European Society of Intensive Care Medicine. Intensive Care Med 1996, 22(7):707–710.

3. Minne L, Abu-Hanna A, de Jonge E: Evaluation of SOFA-based models for predicting mortality in the ICU: A systematic review. Crit Care 2008, 12(6):R161.

4. Minejima E, Wong-Beringer A: Impact of Socioeconomic Status and Race on Sepsis Epidemiology and Outcomes. J Appl Lab Med 2021, 6(1):194–209.

5. Galiatsatos P, Brigham EP, Pietri J, Littleton K, Hwang S, Grant MC, Hansel NN, Chen ES: The effect of community socioeconomic status on sepsis-attributable mortality. J Crit Care 2018, 46:129–133.

6. Galiatsatos P, Follin A, Alghanim F, Sherry M, Sylvester C, Daniel Y, Chanmugam A, Townsend J, Saria S, Kind AJ et al: The Association Between Neighborhood Socioeconomic Disadvantage and Readmissions for Patients Hospitalized With Sepsis. Critical Care Medicine 2020, 48(6):808–814.

7. Chen IY, Szolovits P, Ghassemi M: Can AI Help Reduce Disparities in General Medical and Mental Health Care? AMA J Ethics 2019, 21(2):E167–179.

8. Johnson AE, Stone DJ, Celi LA, Pollard TJ: The MIMIC Code Repository: enabling reproducibility in critical care research. J Am Med Inform Assoc 2018, 25(1):32–39.

9. Johnson A, Pollard T, Blundell J, a-chahin, Gow B erinhong, Schubert M, Dang K, Paris N, shu98 et al: MIT-LCP/mimic-code: MIMIC Code v2.4.0. In., v2.4.0 edn: Zenodo; 2023.

10. Charlson ME, Pompei P, Ales KL, MacKenzie CR: A new method of classifying prognostic comorbidity in longitudinal studies: development and validation. J Chronic Dis 1987, 40(5):373-383.

11. Pedregosa F, Varoquaux G, Gramfort A, Vincent M, Bertrand T, Grisel O, Blondel M, Prettenhofer P, Weiss R, Dubourg V et al: Scikit-learn: Machine Learning in Python. Journal of Machine Learning Research 2011, 12:2825–2830.

12. Patzer RE, Kaji AH, Fong Y: TRIPOD Reporting Guidelines for Diagnostic and Prognostic Studies. JAMA Surg 2021, 156(7):675–676.

13. Raith EP, Udy AA, Bailey M, McGloughlin S, MacIsaac C, Bellomo R, Pilcher DV, Australian, New Zealand Intensive Care Society Centre for O, Resource E: Prognostic Accuracy of the SOFA Score, SIRS Criteria, and qSOFA Score for In-Hospital Mortality Among Adults With Suspected Infection Admitted to the Intensive Care Unit. JAMA 2017, 317(3):290–300.

14. Falcao ALE, Barros AGA, Bezerra AAM, Ferreira NL, Logato CM, Silva FP, do Monte A, Tonella RM, de Figueiredo LC, Moreno R et al: The prognostic accuracy evaluation of SAPS 3, SOFA and APACHE II scores for mortality prediction in the surgical ICU: an external validation study and decision-making analysis. Ann Intensive Care 2019, 9(1):18.

15. de Grooth HJ, Geenen IL, Girbes AR, Vincent JL, Parienti JJ, Oudemans-van Straaten HM: SOFA and mortality endpoints in randomized controlled trials: a systematic review and meta-regression analysis. Crit Care 2017, 21(1):38.

16. Arakawa M, Levy JH, Fujimori K, Kondo K, Iba T: A new SOFA score calculation to improve the predictive performance for mortality in sepsis-associated disseminated intravascular coagulopathy patients. J Crit Care 2021, 64:108–113.

17. Lee HJ, Ko BS, Ryoo SM, Han E, Suh GJ, Choi SH, Chung SP, Lim TH, Kim WY, Kwon WY et al: Modified cardiovascular SOFA score in sepsis: development and internal and external validation. BMC Med 2022, 20(1):263.

18. Liu Y, Gao K, Deng H, Ling T, Lin J, Yu X, Bo X, Zhou J, Gao L, Wang P et al: A time-incorporated SOFA score-based machine learning model for predicting mortality in critically ill patients: A multicenter, real-world study. Int J Med Inform 2022, 163:104776.

19. Levy MM, Fink MP, Marshall JC, Abraham E, Angus D, Cook D, Cohen J, Opal SM, Vincent JL, Ramsay G et al: 2001 SCCM/ESICM/ACCP/ATS/SIS International Sepsis Definitions Conference. Crit Care Med 2003, 31(4):1250–1256.

20. Goldberger AL, Amaral LA, Glass L, Hausdorff JM, Ivanov PC, Mark RG, Mietus JE, Moody GB, Peng CK, Stanley HE: PhysioBank, PhysioToolkit, and PhysioNet: components of a new research resource for complex physiologic signals. Circulation 2000, 101(23):E215–220.

